# A head-mounted Tilted Reality Device for the treatment of pusher syndrome: A usability study

**DOI:** 10.1101/2024.01.22.24301473

**Authors:** Sofia Wöhrstein, Michael Bressler, Lisa Röhrig, Cosima Prahm, Hans-Otto Karnath

**Author notes:** Correspondence should be addressed to: Prof. Dr. Dr. Hans-Otto Karnath, Center of Neurology, University of Tübingen Hoppe-Seyler-Straße 3 D – 72072 Tübingen Germany. These authors contributed equally to this work. **Abbreviations** *ANOVA* – Analysis of Variance; *AT* – Assistive Technology; *HMD* – head-mounted display; *IPD* – interpupillary distance; *IPQ* – iGroup Presence Questionnaire; *PGTQ* – Perception of Game Training Questionnaire; *SSQ* – Simulator Sickness Questionnaire; *SUS* – System Usability Scale; *SVV* – subjective visual vertical; *TRD* – Tilted reality Device; *UX* – user experience; *VFT* – visual feedback training; *VR* – virtual reality.

## Abstract

Pusher syndrome is a disorder of postural control after stroke. Patients show a mismatch in their perception of (almost preserved) visual and (pathologically tilted) postural verticality. In order to reduce this mismatch, we developed a novel head-mounted ‘Tilted Reality Device (TRD)’. It presents patients visual footage of their actual surroundings but tilted to one side rather than upright. We investigated its usability and possible limitations in its use for the treatment of pusher patients in two samples of healthy participants with an average age of 26.4 years and 63.9 years respectively. Individuals from both age groups showed similar levels of tolerance to prolonged exposure to the tilted visual environment for an average of 40.4 minutes while walking around in the hospital. The TRD was found to be comfortable and not frustrating whilst wearing, but somewhat challenging in terms of technical handling, particularly for older participants. At the end of the maximally tolerated exposure time participants of both groups experienced some feelings of discomfort, like dizziness or increased stomach awareness, which disappeared rapidly after terminating TRD exposure. Our TRD appears to be a practical device especially for an older population, like pusher patients. While users must be aware of the possibility of side effects, these should be balanced against the benefits of future use for rehabilitation purposes.

## Introduction

The use of digital media has become increasingly popular to create virtual environments that enable a variety of therapeutic and rehabilitation scenarios for patients suffering from various limitations (Kato, 2010; Primack et al., 2012). So-called ‘games for health‘ include, for example, the rehabilitation of aphasia (Bu et al., 2022), motor control after stroke (Shah et al., 2019; Standen et al., 2017), disorders such as spatial neglect (Huygelier et al., 2020; Knobel et al., 2021; Morse et al., 2020; Stammler et al., 2023), or pusher syndrome (Nestmann et al., 2022).

Based on these developments we developed a novel head-mounted device for the treatment of pusher syndrome. After a unilateral left- or right-hemispheric stroke (Karnath et al., 2000a; Rosenzopf et al., 2023), about 12.5% of hemiparetic patients show a specific disturbance of postural control (Abe et al., 2012; Dai et al., 2022; Pedersen et al., 1996), which has been termed ‘pusher syndrome’ (Davies, 1985; also found as ’contraversive lateropulsion/pushing’ in the literature). The disorder is characterized by a contraversively inclined spontaneous body posture, the use of non-paretic extremities to actively push towards the contralesional side, and active resistance of any external attempts to correct the tilted body posture towards the earth-vertical upright (Davies, 1985; Karnath, 2007; Karnath et al., 2001). Pusher syndrome is based on a faulty perception of onés own body orientation in space (Karnath et al., 2000b). With their eyes closed, pusher patients perceive their body as oriented upright when it is objectively tilted towards the lesion side (Bergmann et al., 2016; Karnath et al., 2000b). In contrast, pusher patients process visual and vestibular information for orientation perception of the visual world (the so-called “subjective visual vertical [SVV]”) almost normally (Johannsen et al., 2006; Karnath et al., 2000b). The resulting mismatch is assumed to function as the pathological mechanism underlying pusher syndrome (Karnath et al., 2000b). While the visual feedback training (VFT; Brötz et al., 2004; Karnath & Brötz, 2003) utilizes conscious use of unimpaired visual-vestibular processing, we here present a novel, non-cognitive approach, the Tilted Reality Device (TRD). It presents the actual, real-time surroundings of a patient via a head-mounted display, captured by a stereo camera. The special feature about the device is that the visual environment is not displayed upright (as is the physical environment) but ipsiversively tilted. This should lead to a reduction of the patient’s mismatch between his/her perception of (almost preserved) visual and (pathologically tilted) postural verticality, and thus enable him/her to (unconsciously) align his/her tilted body posture to earth-gravitational upright.

In the present study, we aimed to determine any possible limitations associated with prospectively using our TRD to treat pusher syndrome and evaluated it in two samples of healthy participants. Since the mean age of patients hospitalized with pusher syndrome after stroke is 68.5 years (Dai et al., 2022), we collected data in a group of older participants and compared it to a younger group of participants to also investigate age-related effects. We were interested in the individual user experience (UX), the user-friendliness and applicability to these two groups wearing the TRD. Beyond, we aimed to explore the amount and extent of symptoms of the so-called ‘cybersickness’, that might potentially occur from manipulated visual input. In principle, symptoms of cybersickness can include disorientation, headache, nausea, dizziness, vertigo, eyestrain and/or difficulty focusing (Bockelman & Lingum, 2017; Rebenitsch & Owen, 2016).

## Methods and Materials

### The Tilted Reality Device (TRD)

Our low-cost head-mounted display (HMD) captures the real environment through a camera, displays it to the user in real-time and thereby allows to feedback the actual visual environment either in upright orientation (as is the physical environment; cf. Fig. 1A), or tilted to one side (Fig. 1B). While the user is wearing the TRD, he/she can walk around and explore the surroundings or simply do whatever he/she wants. A figure of an exemplary participant wearing the TRD had to be removed from the manuscript due to medRxiv restrictions, but can be obtained from the corresponding author on reasonable request.

**Figure 1.**
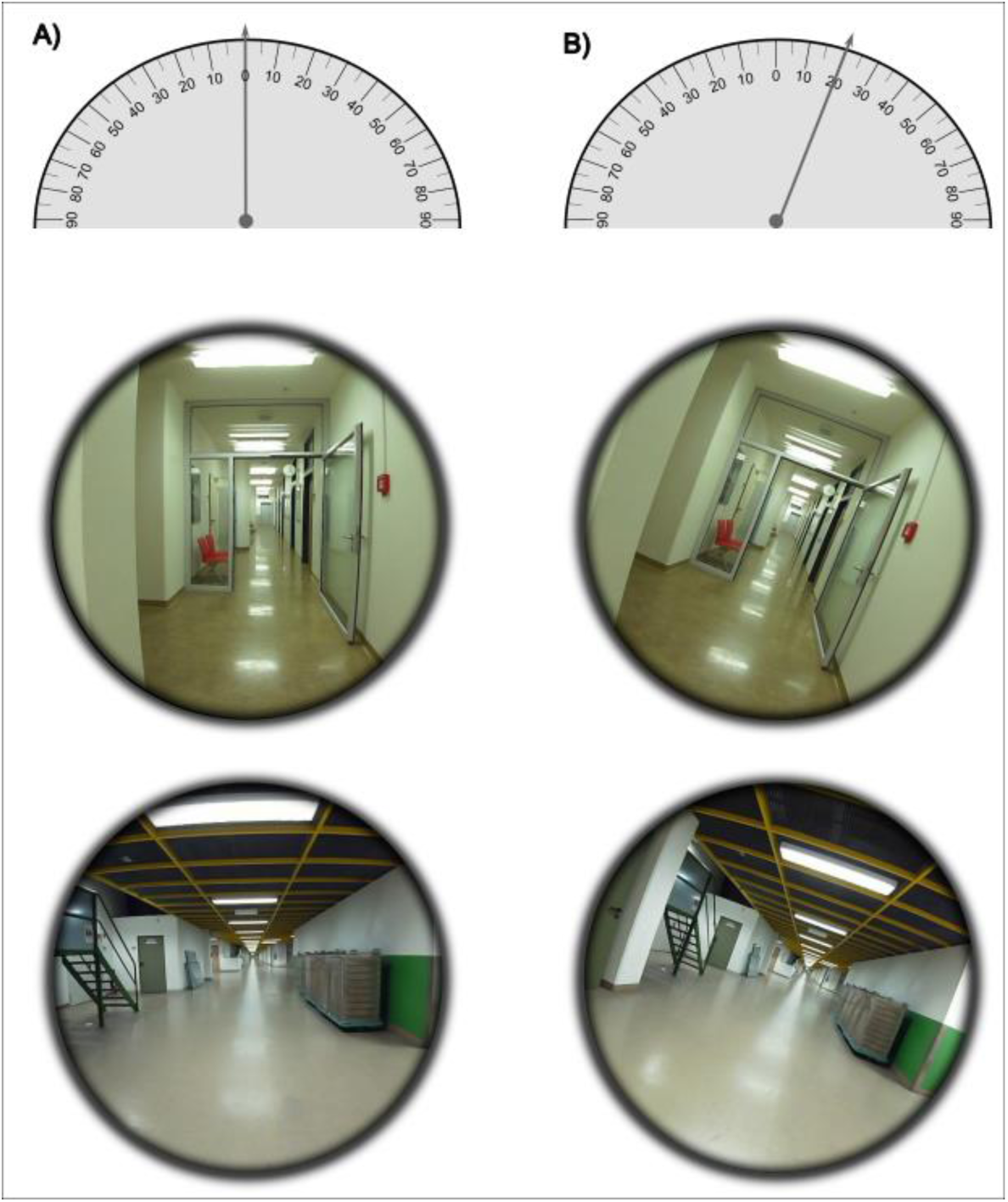
**Tilted Reality Device field of view. A**) Exemplary field of view on the smartphone screen of our Tilted Reality Device (TRD) with no tilt of the visual environment. Depicted are two corridors of different width which were on the route participants took during the experiment while wearing the TRD. **B**) View on the smartphone screen of the TVD with 20° tilt to the right as it was presented in our experiment. The 20° tilt to the right can be read off the goniometer, which tilts a maximum of 90° to the left and right respectively.

**Figure 2.** The TRD. A participant wearing the new Tilted Reality Device (TRD). The figure had to be removed due to medRxiv policy but can be obtained from the corresponding author on request.

The display follows the principle of the Google Cardboard device (Olson et al., 2011). A head-mounted enclosure contains two lenses, in front of which a mobile phone can be attached. Typically, an app on the mobile phone generates the image for both eyes. The rotation of the user’s head is measured by the mobile phone and transferred to the virtual camera. In the present case, instead of a virtual environment, the app displays the video stream from a stereo camera module attached to the front of the display enclosed and connected to the mobile phone. The TRD was built using a generic Cardboard device (manufacturer: bNext, model 8541760985) housing an Android smartphone (Samsung Galaxy S10) with a 6.1-inch display and a resolution of 1440 x 3040 pixels. Instead of using the built-in camera of the phone, we used a 180° wide-angle stereo camera module plugged into the phone (ELP-1080P2CAM-L180). The dual lens camera delivers a high frame rate of 30fps (frames per second) and a 1080p full HD resolution. For the development of the software we used the game development platform Unity (Unity Technologies, 2021). The software application consisted of two components: the mobile Android app, which created the image on the mobile phone in the Cardboard device, and a remote desktop control app that allowed a second person to modify view parameters. The main task of the mobile app is to display the stereoscopic video stream. Due to the functionality of the stereo camera module, it was necessary to enable simultaneous operation of multiple cameras in Unity. For this purpose, an Android OS Unity plugin was developed with Android Studio 4.0 (Google, 2021), which utilized the open source library UVCCamera (Saki, 2017) to communicate with the cameras and integrate the video streams into the resource context of Unity. An OpenGL shading language (GLSL) shader was used to manipulate the video images on a per-pixel basis. To properly adjust the webcam video image to the display, the video stream was horizontally and vertically shifted, and compensation for lens distortion was applied. Furthermore, the video image was circularly shrinked by rendering the outside area black, in order to avoid any visual cues about the extent of the tilt through the edges of the video images. This reduces the field of view on the smartphone screen to 120°. All these parameters could be controlled directly with the mobile app. However, since every change of parameters required removing the display and removing the phone, we created a remote desktop control application to be executed on a computer or laptop. This application connects to the mobile phone via wireless LAN and allows a second person to remotely control all parameters. Finally, the camera module was attached to the display with a goniometer, which allows manual tilting of both lenses together and marking the degree of tilt. The maximum tilt to the left and right was 90° each, which could be read off the goniometer (cf. Fig. 1). The center of rotation was aligned with the participants’ eye level, which equals the mounted cameras height.

### Participants

For our younger sample, a total of 18 neurologically healthy individuals (8 females) between the age of 19 and 39 (*M* = 26.4; *SD* = 4.8) participated in the study. The older group consisted of 18 individuals (9 females) between the age of 55 and 78 (*M* = 63.9; *SD* = 6.4). Participants were recruited through in-house mailing lists. After their arrival, participants were informed about the study procedure and gave their written informed consent in accordance with the Code of Ethics of the World Medical Association (Declaration of Helsinki). After completion of testing, participants were compensated monetarily for their participation. The study was approved by the Ethics Committee of the Medical Faculty of the University of Tübingen, Germany.

### Experimental Procedure

The cameras of the TRD were tilted 20° to the right, creating a corresponding tilt to the right in the visual input image on the screen. Participants put on the device and were then asked to slowly familiarize themselves with the tilted environment by standing up, sitting back down, as well as taking their first steps in the experimental room. This was done until the participant individually felt safe and ready to leave the experimental room. Familiarization lasted about three to four minutes and was included in the total exposure duration. Subsequently, participants walked on a predetermined route through corridors of varying widths (cf. Fig. 1) in the hospital building. Participants were always accompanied by one examiner. They were given the instruction to not alter their head position in response to the tilted environment, which was visually controlled by the experimenter. The maximum exposure duration was set to 45 min, and participants were instructed to verbally report any signs of discomfort in order to terminate the experiment at any time.

### Questionnaires

Right after ending the exposure to the TRD the participants completed a self-compiled questionnaire which consists of ten modified items taken from the following questionnaires: the Perception of Game Training Questionnaire (PGTQ; Boot et al., 2013), the System Usability Scale (SUS; Brooke, 1996), and the iGroup Presence Questionnaire (IPQ; Schubert, 2003; http://www.igroup.org/pq/ipq/ipq_german.htm) to assess the subjective user experience (UX), usability, and user-friendliness of our TRD. Additionally, the SSQ (cf. paragraph after next) was applied twice to obtain pre and post TRD exposure scores regarding potential side effects. The items from all questionnaires were translated to German.

Three items were taken from the PGTQ to assess how challenging, enjoyable and frustrating the participants experienced the exposure to the tilted vision. The items were rated on a 5-point Likert scale ranging from “strongly agree” (5) to “strongly disagree” (1) with 3 as a neutral midpoint. Since each item represents a different aspect of the individual UX, each item was considered separately, and no mean score was computed. The SUS was used to evaluate the handling of the TRD. Five out of the original ten items were included in our compiled questionnaire and were rated on a 5-point Likert scale ranging from “strongly agree” (5) to “strongly disagree” (1), again using 3 as a neutral midpoint. As we did not use the entire SUS, our analysis differed from the details in the original publication, and we computed an overall mean score which represents the difficulty or rather simplicity of handling. Two of the five items (items 6 and 8 in our questionnaire) were coded negative, so they had to be recoded before further analysis. The IPQ depicts the sense of presence in a virtual environment, that is the sense of being there (Schubert, 2003). Since we do not use a virtual environment but a tilted, real one, we only included two items from the IPQ which measure the realness of the new environment. This way participants could judge how ‘real’ the tilted vision in our TRD is. Both items were rated on a 5-point Likert scale; one of them ranging from “not at all” (1) to “entirely” (5) and the other one from “not real at all” (1) to “perfectly real” (5). Again, a score of 3 served as a neutral midpoint. We computed a mean score across both items. For all these items or questionnaires, we also tested if there were statistical differences between our younger and older group of participants.

Moreover, a slightly modified version of the Simulator Sickness Questionnaire (SSQ; Kennedy et al., 1993) was applied at the beginning and after the TRD exposure. The aim was to assess possible side effects of being exposed to a tilted environment for a longer time duration. The following 14 symptoms were queried by the experimenter as soon as participants started to wear the TRD and directly after taking it off: general discomfort, fatigue, headache, eyestrain, difficulty focusing, increased salivation, sweating, nausea, difficulty concentrating, fullness of head, dizzy, vertigo, stomach awareness, and burping. Symptoms were verbally rated on a 4-point Likert scale ranging from 0 to 3 (none, slightly, moderately, severely).

### Statistical analysis

Since we aimed to investigate the effects of the tilted environment exposure on cybersickness ratings, including the possibility that there were no differences between the pre- and post-ratings, we chose a Bayesian statistical approach (cf. Huygelier et al., 2020). Our within-subjects ANOVA model included all 14 SSQ items as repeated measures within participants as well as the group assignment (young vs. old group) as an interacting effect with the tilted environment exposure. This was done using the *BayesFactor* package (Rouder et al., 2012; Rouder et al., 2009) in *R Studio* (Posit Team, 2022).

## Results

The targeted exposure duration of 45 minutes was not reached by every participant due to discomfort and consequently earlier termination. In the sample of younger participants, the exposure to tilted vision lasted on average 40.6 minutes (*SD* = 6.0) and ranged between 29 and 45 minutes. The average duration in the group of older participants was 40.3 minutes (*SD* = 10.4) and ranged between 12 and 45 minutes. The average exposure duration did not differ significantly between the two groups (*t*(27.21) = -0.12, *p* = .907), indicating that individuals from both age groups exhibited a similar level of tolerance for prolonged exposure to the tilted visual environment. The higher variance in the exposure duration of the elder group was especially due to one participant who felt sick already after 12 min (cf. paragraph after next).

On average, the experience of frustration with our TRD was statistically equally low in both groups (answers ranging between *disagreement* and *neutral*), however they perceived the exposure as somewhat challenging (Tab. 1). This suggests that although wearing the TRD was challenging for some older participants, it did not lead to significant frustration. Significant differences were found for the comparison of the two age groups in the PGTQ for the item assessing how enjoyable the participants experienced the exposure to the tilted environment (*t*(28.20) = -2.60, *p* = .015), as well as for the SUS (*t*(24.28) = -2.23, *p* = .036) which surveys the difficulty or rather simplicity of handling of the TRD (for all results cf. Tab. 1). In both measures the younger group obtained higher scores, that is, they rated the usability higher than older participants and also had more fun during the exposure (Fig. 3; for a detailed overview of the individual scores for each item of the questionnaires see supplementary Fig. S1 to S3). The scores in the IPQ, measuring the feeling of realness of the new environment with the TRD, ranged around the neutral midpoint. This suggests that our participants perceived the tilted real-life environment as realistic but did not experience a sense of immersion in a new reality.

**Figure 3.**
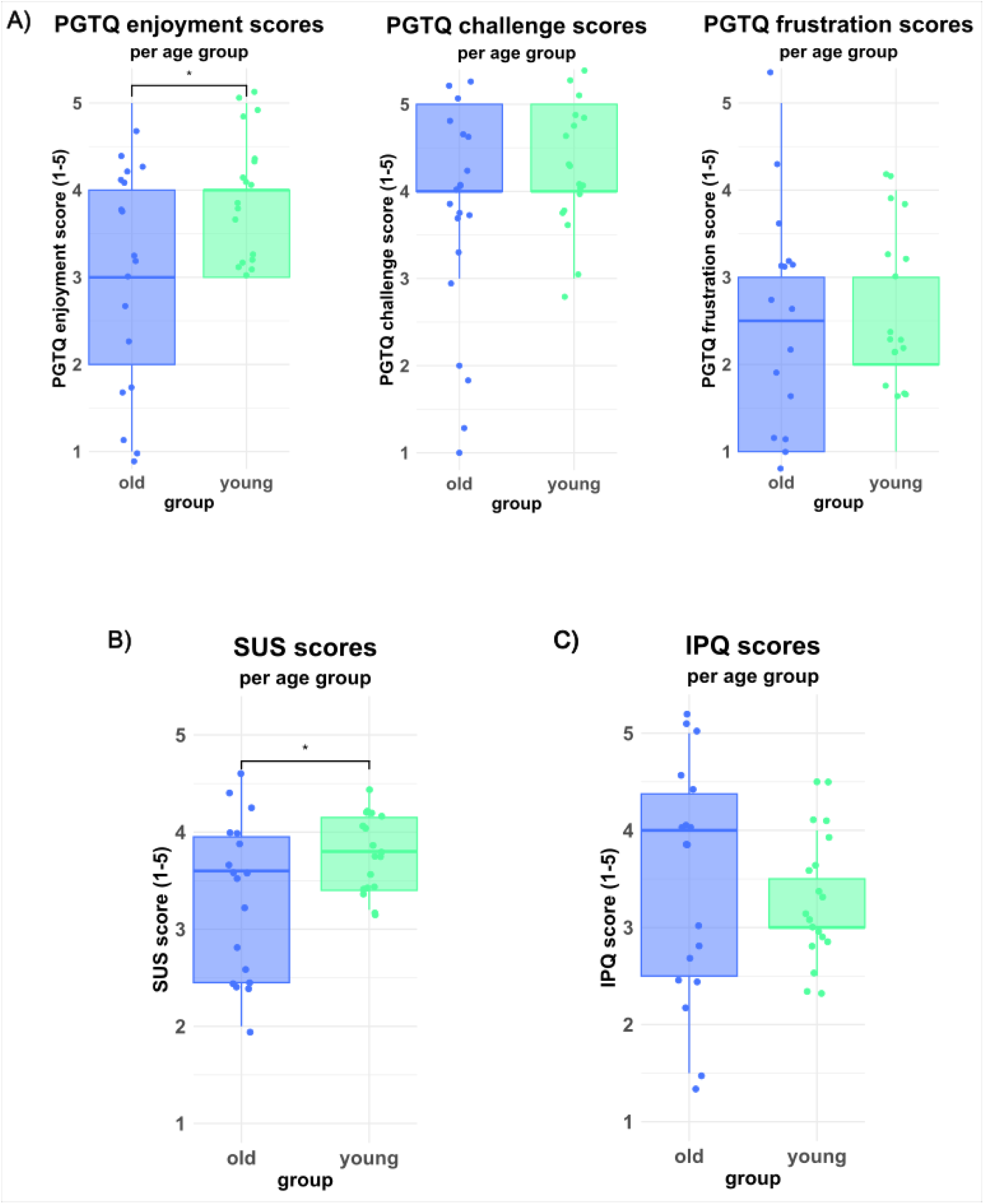
Questionnaire scores. Questionnaire scores of the healthy older and younger adults evaluating the use of the Tilted Reality Device (TRD). Illustrated are boxplots with the median and quartile ranges of the respective group score. Individual values were jittered for better visibility of all measured values. **A**) Perception of Game Training Questionnaire (PGTQ) on the dimensions “enjoyment”, “challenge” and “frustration”, measured on a 5-point Likert scale (1 = “strongly disagree”, 5 = “strongly agree”). Illustrated are mean response values for the two age groups. Only the enjoyment scores showed a significant difference (α = 0.05) between the young and the older experimental group. **B**) System Usability Scale (SUS) measuring the simplicity of handling the device. Depicted are the significantly different mean response values for the two age groups on a 5-point Likert scale from 1 = “strongly disagree” to 5 = “strongly agree”. **C**) iGroup Presence Questionnaire (IPQ) measuring the sense of presence in the new environment. Portrayed are the mean response values of both age groups, rated on a 5-point Likert scale from 1 = “not real at all” to 5 = “perfectly real”.

**Table 1.**
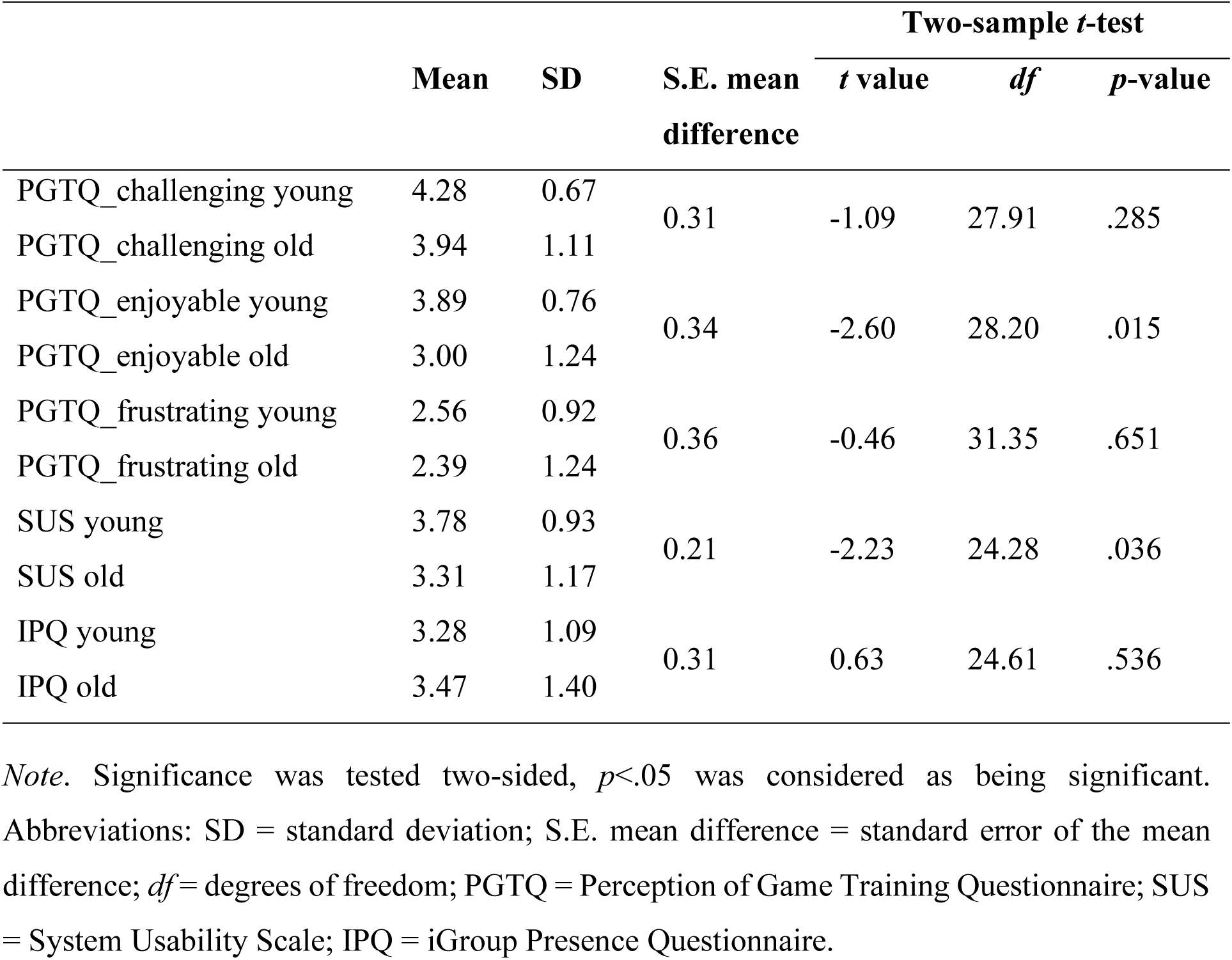
Statistical results comparing the young and the older group of healthy participants on the different questionnaires.

When looking at possible side effects of being exposed to the tilted environment, assessed with the SSQ, the age groups did not differ significantly in their ratings. This was the case for the pre-exposure ratings (*Myoung* = 0.17, *SDyoung* = 0.12; *Mold* = 0.10, *SDold* = 0.14; *t*(33.24) = -1.69, *p* = .099) as well as the post-exposure ratings (*Myoung* = 0.79, *SDyoung* = 0.71; *Mold* = 0.61, *SDold* = 0.68; *t*(33.91) = -0.81, *p* = .424). The average ratings in both groups were below 1, which indicates only a slight sensation of a specific symptom. A detailed overview of all 14 SSQ symptom pre- and post-exposure scores is given in Figure 4. The Bayesian model analyzing the SSQ before and after the exposure to tilted vision showed no effect of groups (young vs. old; BF10 = -1.09) but strong evidence for an effect of intervention (BF10 = 36.11). The latter indicates that the experience of cybersickness symptoms, as measured with the SSQ, was stronger after the average 40.4 min TRD exposure than at the beginning.

**Figure 4.**
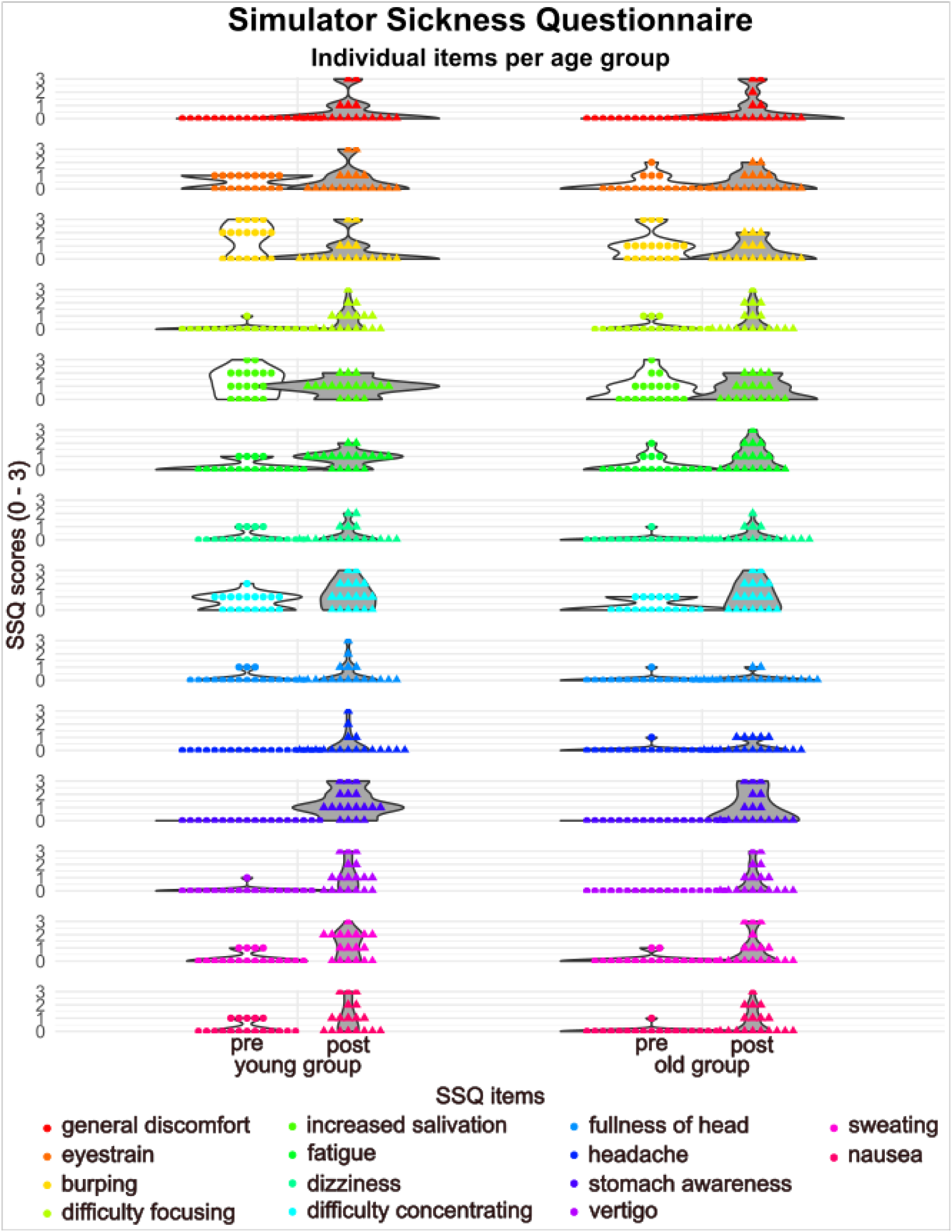
Cybersickness ratings. Single item ratings in the Simulator Sickness Questionnaire (SSQ) at the beginning (pre) and after ending (post) the exposure to the Tilted Reality Device (TRD) separate for both age groups. The violin plots illustrate the distribution of group ratings for each item. The items were rated on a 4-point Likert scale ranging from 0 to 3 (0 - none, 1 - slightly, 2 - moderately, 3 - severely).

One participant of the elderly group experienced sickness already 12 minutes after commencing the TRD exposure, leading to discontinuation of the experiment. She primarily complained of nausea and heightened stomach awareness, subsequently resulting in vomiting. The participant verbally reported that she felt better and nearly back to normal 10 minutes after emesis. No other participant experienced such severe side effects. Two other participants in the elder group terminated the experiment early (one after 18 minutes, the other after 25 minutes) due to feelings of discomfort (a “funny feeling in the stomach”). In the younger group, six participants terminated the experiment before the maximum duration of 45 minutes, with termination times of 29, 30, 31, 33, 36, and 39 minutes respectively. Participants reported either a discomforting sensation in their stomach or dizziness as reasons for discontinuing their exposure to the tilted environment wearing the TRD.

## Discussion

The present study aimed to develop and assess the usability, practicality, as well as possible usage restrictions of a novel Tilted Reality Device (TRD) designed for the therapy of pusher syndrome. An important aspect under examination was the duration of exposure that healthy participants tolerate while wearing the TRD. We found that, on average, participants tolerated approximately 40.4 minutes of exposure, with a notable range of individual durations varying from 12 to 45 minutes across the entire sample, irrespective of age. We observed no significant difference in average exposure duration between the younger and older participant groups. At the end of the maximally tolerated exposure time participants of both groups experienced some feelings of discomfort, like dizziness or increased stomach awareness, but these feelings remained below a score of one (representing *slightly*) and disappeared rapidly after terminating TRD exposure. There were no significant differences between the age groups in their pre-exposure and post-exposure ratings, indicating that the two groups exhibited similar levels of specific symptoms related to cybersickness. This indicates that our TRD is a tool which − also in an older population like pusher patients − is tolerated on average for up to ∼40 minutes before the occurrence of significant side effects.

Our study also encompassed an assessment of the participants’ user experience (UX) and the usability as well as practicality of the TRD. We observed a significant difference between the two age groups in their ratings of enjoyment during the tilted vision exposure, as well as their perception of the TRD’s usability. Specifically, the younger group expressed higher levels of enjoyment and rated the TRD as more user-friendly compared to their older counterparts. This discrepancy suggests that younger individuals found the TRD experience to be more engaging and approachable, possibly due to their more natural socialization with digital technologies. However, the experience of frustration with our TRD was statistically equally low in both groups, suggesting that although wearing the TRD was challenging for some older participants, it did not lead to significant frustration, which is positive in terms of future therapeutic application.

This study also examined the perceived authenticity of the tilted visual environment. The results indicated that participants’ scores hovered around the neutral midpoint, suggesting that the realism of the new environment with the TRD was neither strongly affirmed nor denied. This can be interpreted as a sign that our participants did not perceive the tilted environment as particularly changed in contrast to their perception without the TRD. As mentioned above, our future goal is to use the TRD for the treatment of pusher syndrome, where patients are only expected to unconsciously process the tilted environment. The present finding supports the idea of such an unconscious processing mechanism when using the TRD, since the visual manipulation can remain undetected to a certain extent. This principle aims at the aforementioned idea that wearing the TRD does not necessarily have to be linked to a conscious, active therapeutic action for the patient but could also represent a possibly unconscious support for the pusher patient. This could be a significant help, especially in the first period after the stroke, when patients are most affected and cannot sit or stand independently. From this point of view, our TRD could be considered a so-called ‘Assistive Technology (AT)’ that supports existing therapeutic options such as physiotherapy and the visual feedback training (Brötz et al., 2004; Karnath & Brötz, 2003). AT is generally understood to be a type of technological intervention for rehabilitation purposes that serves people with acquired impairments and disabilities. It provides extrinsic support and aims to target the remaining functional abilities of the person affected (LoPresti et al., 2004).

Overall, our newly developed TRD is well suited for its intended use in pusher patients. In this usability assessment study, only one participant (i.e. 3% of the total sample) experienced severe side effects due to the exposure to a tilted environment. This can be explained by the aforementioned phenomenon of cybersickness. The two most prominent theories regarding the causes of cybersickness are the sensory mismatch theory (Reason, 1978) and the postural instability theory (Riccio & Stoffregen, 1991). The sensory mismatch theory states that the symptoms of cybersickness occur due to different perceptions of environmental stimuli by different senses (Reason, 1978; Rebenitsch & Owen, 2016). In our case this translates to the following situation: the visual system perceives a misaligned environment, while the vestibular system does not detect any anti-gravitational tilt since the participants are standing and walking in their typical vertical posture. Another cause for the sensory mismatch presumably lies in the stereo camera used in the TRD, which has a maximum frame rate of 30 images per second. Fast head movements thus can lead to the perception of a slight latency of the projected video stream and thus also can cause cybersickness (Palmisano et al., 2020). The second hypothesis, the postural instability theory, claims that cybersickness arises from the failure to sustain the appropriate posture for the processing of a particular environmental stimulus (Rebenitsch & Owen, 2016; Riccio & Stoffregen, 1991). Again, for our participants this implies that cybersickness may develop due to their inability to maintain a vertical body tilt of 20° while moving in accordance with the presented visual stimulus. Be that as it may, our findings suggest that supporting the rehabilitative process of stroke patients affected by pusher syndrome through the use of the TRD could become reasonable, as all but one (i.e. 97%) of our participants showed good tolerance of the TRD.

Although our device does not use a virtual but the real environment, a comparison with the experiences gained when using virtual reality (VR) technology may nevertheless be useful. We observed that both of our age groups tolerated wearing the new TRD for an average duration of about 40 minutes while walking around in our hospital before they experienced some feelings of discomfort. It is known that the duration of exposure to a *virtual* environment also has an impact on cybersickness, with a longer exposure leading to increased cybersickness ratings (Kennedy et al., 2000). Although this finding is considered certain, there is little consensus on the length of time that should be considered the upper limit of exposure duration. This is due to the fact that cybersickness is subject to eminently individual influencing factors like, for example, sex (Munafo et al., 2017; Stanney et al., 2020), cybersickness history (Stanney et al., 2020; Stanney et al., 2003), postural (in-)stability (Arcioni et al., 2019; Risi & Palmisano, 2019), and interpupillary distance (IPD; Fulvio et al., 2021; Stanney et al., 2020), i.e. the distance between the pupils relevant for stereo vision. In addition, it is known about the use of VR technology that repeated exposures to virtual environments are known to significantly reduce cybersickness scores due to adaptation effects (Kennedy et al., 2000). After only a few exposures (sometimes already during the second exposure) users become habituated such that ratings in cybersickness scales drop drastically (Dużmańska et al., 2018; Gavgani et al., 2017; Hill & Howarth, 2000; Howarth & Hodder, 2008; Risi & Palmisano, 2019). This observation with the use of VR technologies could indicate that the repeated use of our TRD, e.g. during regular therapy sessions, could prove to be even more advantageous for patients in terms of tolerability than was measured here for a single exposure. Nevertheless, future users should be aware that some individuals may occasionally not tolerate the application so well. In this case, however, we observed that those affected recovered quickly from the unpleasant sensations after removing the TRD.

After testing the applicability of the new device in the present study, we are confident that the use of the TRD in pusher patients will actually have therapeutic success. The reason for this is that our group has already been able to show in a single case study, albeit with using VR technology, that the principle used here, namely presenting the patient with the visual environment in a tilted state, was able to reduce pusher symptoms (Nestmann et al., 2022). The authors used a three-dimensional virtual environment (a scene of a beach with a footbridge) that could be explored by wearing a HMD. The authors manipulated the 3D visual input in the

VR setup by tilting the horizon of the visual scene presented to the patient, in order to reduce the mismatch between the different modalities of verticality perception in pusher syndrome. In contrast to such an artificial VR scenery, however, our TRD offers real-time vision of the actual, authentic environment in which the user is located, although tilted sideways. In this way, the user can see what he/she would normally see and can actively move around and explore his/her real surroundings. The user is not forced to sit still on a chair while using the device but can move freely. A related device was reported by Greenberg et al. (2017), but it had several shortcomings, leading the authors to conclude that they were unable to develop a functional device due to limitations in the hardware as well as software components. Specifically, the device lacked a stereoscopic image, the image could not be tilted properly, did not match the entire view area of the video stream image, visual cues indicating the actual tilt to the user, such as the edge of the video image, were clearly visible, and eye positioning was incorrect. In contrast, our new TRD has taken all these aspects into account from the outset to develop a technically high-quality device.

## Conclusion

Our study demonstrates the feasibility of the newly developed Tilted Reality Device (TRD). The findings highlight the user friendliness and practicality of the TRD, as well as its maximally tolerated exposure time to the tilted visual environment of ∼40 minutes, which also applies to an older population such as pusher patients. However, future users should be aware of the possibility of experiencing symptoms of cybersickness. Perhaps a sensible use of the device should initially not exceed a duration of 30 minutes per single application in order to counteract the occurrence of side effects. To achieve an even higher level of user-friendliness, our TRD could be enhanced, for example, by the use of a stereo camera with a higher frame rate to further reduce the latency of the video image and thus reduce the occurrence of side effects to a greater extent. All in all, the present results demonstrate the potential of the TRD as a viable tool for rehabilitation purposes in stroke patients affected by pusher syndrome.

## Data Availability

All data produced in the present study are available upon reasonable request to the authors.

## Disclosure

The authors report no competing interests.

## Notes

### Competing Interest Statement

The authors have declared no competing interest.

### Funding Statement

This study did not receive any external funding.

### Author Declarations

The ethics committee at the medical faculty of the Eberhard-Karls-University at Tuebingen University Hospital gave ethical approval for this work.

## References

Abe, H., Kondo, T., Oouchida, Y., Suzukamo, Y., Fujiwara, S., & Izumi, S.-I. (2012). Prevalence and length of recovery of pusher syndrome based on cerebral hemispheric lesion side in patients with acute stroke. Stroke, 43(6), 1654–1656. 10.1161/STROKEAHA.111.638379

Arcioni, B., Palmisano, S., Apthorp, D., & Kim, J. (2019). Postural stability predicts the likelihood of cybersickness in active HMD-based virtual reality. Displays, 58, 3–11. 10.1016/j.displa.2018.07.001

Bergmann, J., Krewer, C., Selge, C., Müller, F., & Jahn, K. (2016). The subjective postural vertical determined in patients with pusher behavior during standing. Topics in Stroke Rehabilitation, 23(3), 184–190. 10.1080/10749357.2015.1135591

Bockelman, P., & Lingum, D. (2017). Factors of cybersickness. In C. Stephanidis (Ed.), HCI International 2017 - Posters’ Extended Abstracts. Communications in Computer and Information Science. (Vol. 714, pp. 3–8). Springer International Publishing. 10.1007/978-3-319-58753-0_1

Boot, W. R., Champion, M., Blakely, D. P., Wright, T., Souders, D., & Charness, N. (2013). Video games as a means to reduce age-related cognitive decline: attitudes, compliance, and effectiveness. Frontiers in Psychology, 4, 31. 10.3389/fpsyg.2013.00031

Brooke, J. (1996). SUS: a ’quick and dirty’ usability scale. In P. W. Jordan, B. Thomas, B. A. Weerdmeester, & I. L. McClelland (Eds.), Usability Evaluation in Industry (Vol. 189). Taylor & Francis Ltd.

Brötz, D., Johannsen, L., & Karnath, H. O. (2004). Time course of ‘pusher syndrome’ under visual feedback treatment. Physiotherapy Research International, 9(3), 138–143. 10.1002/pri.314

Bu, X., Ng, P. H., Tong, Y., Chen, P. Q., Fan, R., Tang, Q., Cheng, Q., Li, S., Cheng, A. S., & Liu, X. (2022). A mobile-based virtual reality speech rehabilitation app for patients with aphasia after stroke: development and pilot usability study. JMIR Serious Games, 10(2), e30196. 10.2196/30196

Dai, S., Lemaire, C., Piscicelli, C., & Pérennou, D. (2022). Lateropulsion Prevalence After Stroke: A Systematic Review and Meta-analysis. Neurology (1526-632X (Electronic)). 10.1212/WNL.0000000000200010

Davies, P. M. (1985). Steps to Follow. A Guide to the Treatment of Adult Hemiplegia (1 ed.). Springer Berlin, Heidelberg.

Dużmańska, N., Strojny, P., & Strojny, A. (2018). Can Simulator Sickness be Avoided? A Review on Temporal Aspects of Simulator Sickness. Frontiers in Psychology, 9, 2132. 10.3389/fpsyg.2018.02132

Fulvio, J. M., Ji, M., & Rokers, B. (2021, 2021/05/01/). Variations in visual sensitivity predict motion sickness in virtual reality. Entertainment Computing, 38, 100423. 10.1016/j.entcom.2021.100423

Gavgani, A. M., Nesbitt, K. V., Blackmore, K. L., & Nalivaiko, E. (2017). Profiling subjective symptoms and autonomic changes associated with cybersickness. Autonomic Neuroscience: Basic and Clinical, 203, 41–50. 10.1016/j.autneu.2016.12.004

Google. (2021). Android Studio [Integrated Development Environment]. https://developer.android.com/studio

Greenberg, A., Hancock, F., & Patino, F. (2017). Mobile Augmented Reality as Rehabilitation for Lateropulsion. Proceedings of Student-Faculty Research Day, Pace University, New York, USA. https://api.semanticscholar.org/CorpusID:28232153.

Hill, K. J., & Howarth, P. A. (2000). Habituation to the side effects of immersion in a virtual environment. Displays, 21(1), 25–30. 10.1016/S0141-9382(00)00029-9

Howarth, P. A., & Hodder, S. G. (2008). Characteristics of habituation to motion in a virtual environment. Displays, 29(2), 117–123. 10.1016/j.displa.2007.09.009

Huygelier, H., Schraepen, B., Lafosse, C., Vaes, N., Schillebeeckx, F., Michiels, K., Note, E., Vanden Abeele, V., van Ee, R., & Gillebert, C. R. (2020). An immersive virtual reality game to train spatial attention orientation after stroke: A feasibility study. Applied Neuropsychology: Adult, 1-21. 10.1080/23279095.2020.1821030

Johannsen, L., Fruhmann Berger, M., & Karnath, H.-O. (2006). Subjective visual vertical (SVV) determined in a representative sample of 15 patients with pusher syndrome. Journal of Neurology, 253(10), 1367–1369. 10.1007/s00415-006-0216-x

Karnath, H.-O. (2007). Pusher syndrome – a frequent but little-known disturbance of body orientation perception. Journal of Neurology, 254(4), 415–424. 10.1007/s00415-006-0341-6

Karnath, H.-O., & Brötz, D. (2003). Understanding and Treating “Pusher Syndrome”. Physical Therapy, 83(12), 1119–1125. 10.1093/ptj/83.12.1119

Karnath, H.-O., Brötz, D., & Götz, A. (2001). Klinik, Ursache und Therapie der Pusher-Symptomatik. Der Nervenarzt, 72(2), 86–92. 10.1007/s001150050719

Karnath, H.-O., Ferber, S., & Dichgans, J. (2000a). The neural representation of postural control in humans. Proceedings of the National Academy of Sciences, 97(25), 13931–13936. 10.1073/pnas.240279997

Karnath, H.-O., Ferber, S., & Dichgans, J. (2000b). The origin of contraversive pushing: Evidence for a second graviceptive system in humans. Neurology, 55(9), 1298–1304. 10.1212/WNL.55.9.1298

Kato, P. M. (2010). Video Games in Health Care: Closing the gap. Review of General Psychology, 14(2), 113–121. 10.1037/a0019441

Kennedy, R. S., Lane, N. E., Berbaum, K. S., & Lilienthal, M. G. (1993). Simulator Sickness Questionnaire: An Enhanced Method for Quantifying Simulator Sickness. The International Journal of Aviation Psychology, 3(3), 203–220. 10.1207/s15327108ijap0303_3

Kennedy, R. S., Stanney, K. M., & Dunlap, W. P. (2000). Duration and Exposure to Virtual Environments: Sickness Curves During and Across Sessions. Presence: Teleoperators & Virtual Environments, 9(5), 463–472. 10.1162/105474600566952

Knobel, S. E. J., Kaufmann, B. C., Gerber, S. M., Urwyler, P., Cazzoli, D., Müri, R. M., Nef, T., & Nyffeler, T. (2021). Development of a Search Task using Immersive Virtual Reality: Proof-of-Concept Study. JMIR Serious Games, 9(3), e29182. 10.2196/29182

LoPresti, E. F., Mihailidis, A., & Kirsch, N. (2004). Assistive Technology for Cognitive Rehabilitation: State of the Art. Neuropsychological Rehabilitation, 14(1-2), 5–39. 10.1080/09602010343000101

Morse, H., Biggart, L., Pomeroy, V., & Rossit, S. (2020). Exploring perspectives from stroke survivors, carers and clinicians on virtual reality as a precursor to using telerehabilitation for spatial neglect post-stroke. Neuropsychological Rehabilitation, 1-25. 10.1080/09602011.2020.1819827

Munafo, J., Diedrick, M., & Stoffregen, T. A. (2017). The virtual reality head-mounted display Oculus Rift induces motion sickness and is sexist in its effects. Experimental Brain Research, 235(3), 889–901. 10.1007/s00221-016-4846-7

Nestmann, S., Röhrig, L., Müller, B., Ilg, W., & Karnath, H.-O. (2022). Tilted 3D visual scenes influence lateropulsion: A single case study of pusher syndrome. Journal of Clinical and Experimental Neuropsychology, 44(7), 478–486. 10.1080/13803395.2022.2121382

Olson, J. L., Krum, D. M., Suma, E. A., & Bolas, M. (2011). A design for a smartphone-based head mounted display. 2011 IEEE Virtual Reality Conference, Singapore.

Palmisano, S., Allison, R. S., & Kim, J. (2020). Cybersickness in Head-Mounted Displays is Caused by Differences in the User’s Virtual and Physical Head Pose. Frontiers in Virtual Reality, 1, 587698. 10.3389/frvir.2020.587698

Pedersen, P. M., Wandel, A., Jørgensen, H. S., Nakayama, H., Raaschou, H. O., & Olsen, T. S. (1996). Ipsilateral pushing in stroke: Incidence, relation to neuropsychological symptoms, and impact on rehabilitation. The Copenhagen stroke study. Archives of Physical Medicine and Rehabilitation, 77(1), 25–28. 10.1016/S0003-9993(96)90215-4

Posit Team. (2022). RStudio: Integrated Development Environment for R. In (Version 2022.12.0.353) Posit Software, PBC. http://www.posit.co/.

Primack, B. A., Carroll, M. V., McNamara, M., Klem, M. L., King, B., Rich, M., Chan, C. W., & Nayak, S. (2012). Role of Video Games in Improving Health-Related Outcomes: A Systematic Review. American Journal of Preventive Medicine, 42(6), 630–638. 10.1016/j.amepre.2012.02.023

Reason, J. T. (1978). Motion Sickness Adaptation: A Neural Mismatch Model. Journal of the Royal Society of Medicine, 71(11), 819–829. 10.1177/014107687807101109

Rebenitsch, L., & Owen, C. (2016). Review on cybersickness in applications and visual displays. Virtual Reality, 20(2), 101–125. 10.1007/s10055-016-0285-9

Riccio, G. E., & Stoffregen, T. A. (1991). An ecological Theory of Motion Sickness and Postural Instability. Ecological Psychology, 3(3), 195–240. 10.1207/s15326969eco0303_2

Risi, D., & Palmisano, S. (2019). Effects of postural stability, active control, exposure duration and repeated exposures on HMD induced cybersickness. Displays, 60, 9–17. 10.1016/j.displa.2019.08.003

Rosenzopf, H., Klingbeil, J., Wawrzyniak, M., Röhrig, L., Sperber, C., Saur, D., & Karnath, H.-O. (2023). Thalamocortical disconnection involved in pusher syndrome. Brain, 00, 1–14. 10.1093/brain/awad096

Rouder, J. N., Morey, R. D., Speckman, P. L., & Province, J. M. (2012). Default Bayes factors for ANOVA designs. Journal of Mathematical Psychology, 56(5), 356–374. 10.1016/j.jmp.2012.08.001

Rouder, J. N., Speckman, P. L., Sun, D., Morey, R. D., & Iverson, G. (2009). Bayesian t tests for accepting and rejecting the null hypothesis. Psychonomic Bulletin & Review, 16, 225–237. 10.3758/PBR.16.2.225

Saki, T. (2017). UVCCamera. In https://github.com/saki4510t/UVCCamera

Schubert, T. W. (2003). The sense of presence in virtual environments: A three-component scale measuring spatial presence, involvement, and realness. Zeitschrift für Medienpsychologie, 15(2), 69–71. 10.1026//1617-6383.15.2.69

Shah, V., Cuen, M., McDaniel, T., & Tadayon, R. (2019). A Rhythm-Based Serious Game for Fine Motor Rehabilitation Using Leap Motion. 2019 58th Annual Conference of the Society of Instrument and Control Engineers of Japan (SICE), Hiroshima, Japan.

Stammler, B., Flammer, K., Schuster, T., Lambert, M., & Karnath, H.-O. (2023). Negami: An Augmented Reality App for the Treatment of Spatial Neglect After Stroke. JMIR Serious Games, 11, e40651. 10.2196/40651

Standen, P., Threapleton, K., Richardson, A., Connell, L., Brown, D., Battersby, S., Platts, F., & Burton, A. (2017). A low cost virtual reality system for home based rehabilitation of the arm following stroke: a randomised controlled feasibility trial. Clinical Rehabilitation, 31(3), 340–350. 10.1177/0269215516640320

Stanney, K., Fidopiastis, C., & Foster, L. (2020). Virtual Reality Is Sexist: But It Does Not Have to Be. Frontiers in Robotics and AI, 7(2296-9144 (Electronic)). 10.3389/frobt.2020.00004

Stanney, K. M., Hale, K. S., Nahmens, I., & Kennedy, R. S. (2003). What to Expect from Immersive Virtual Environment Exposure: Influences of Gender, Body Mass Index, and Past Experience. Human Factors, 45(3), 504–520. 10.1518/hfes.45.3.504.27254

Unity Technologies. (2021). Unity 2021 [Software]. https://unity.com

