## Supplementary Figures for "A head-mounted Tilted Reality Device for the treatment of pusher syndrome: A usability study"

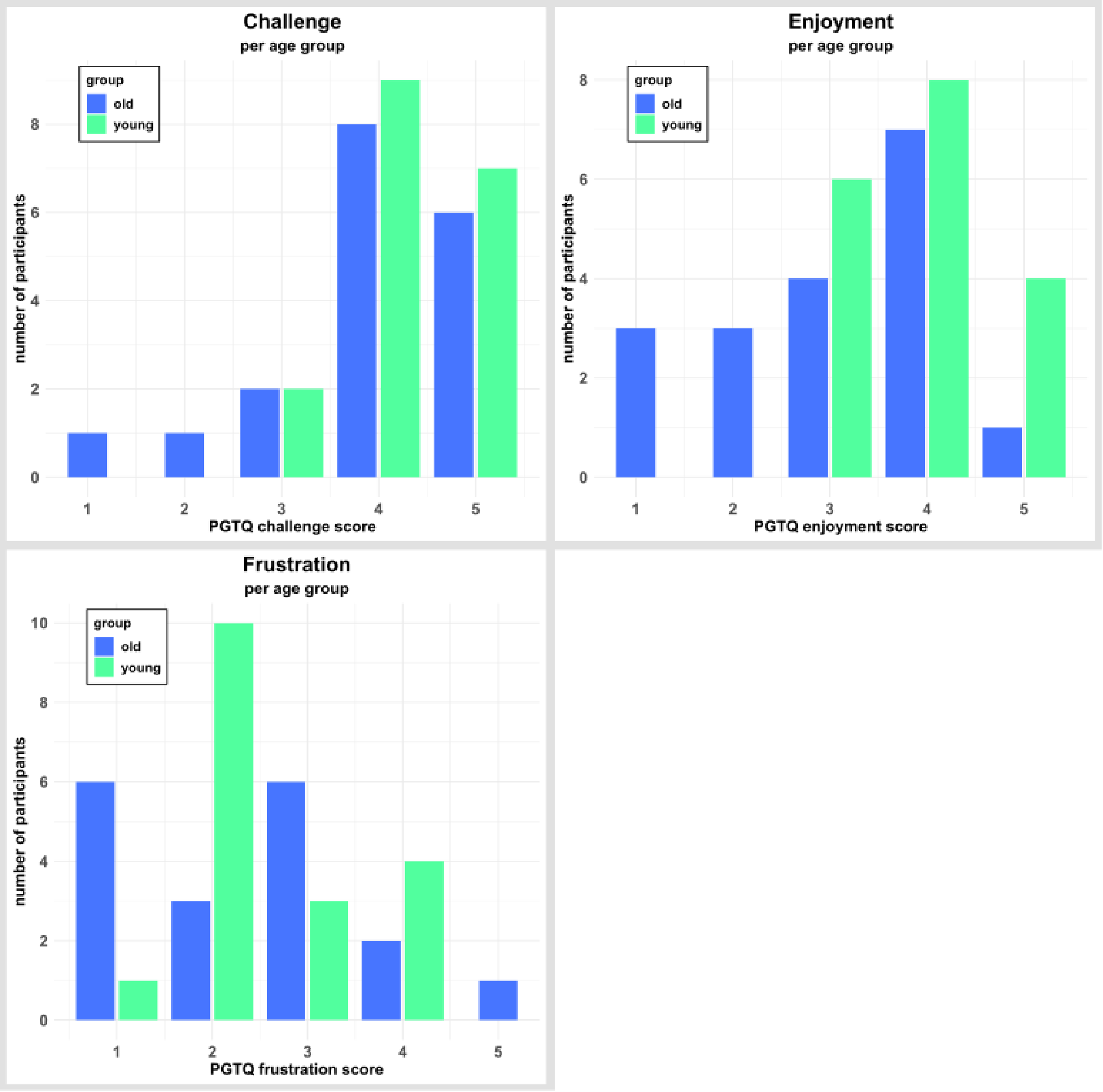


**Figure S1. Individual item scores of the Perception of Game Training Questionnaire (PGTQ).** Shown are scores on the dimensions challenge, enjoyment, and frustration. The items were rated on a 5-point Likert scale (1 = "strongly disagree", 5 = "strongly agree"). Bar plots represent the number of participants per possible item score, separated by age group.

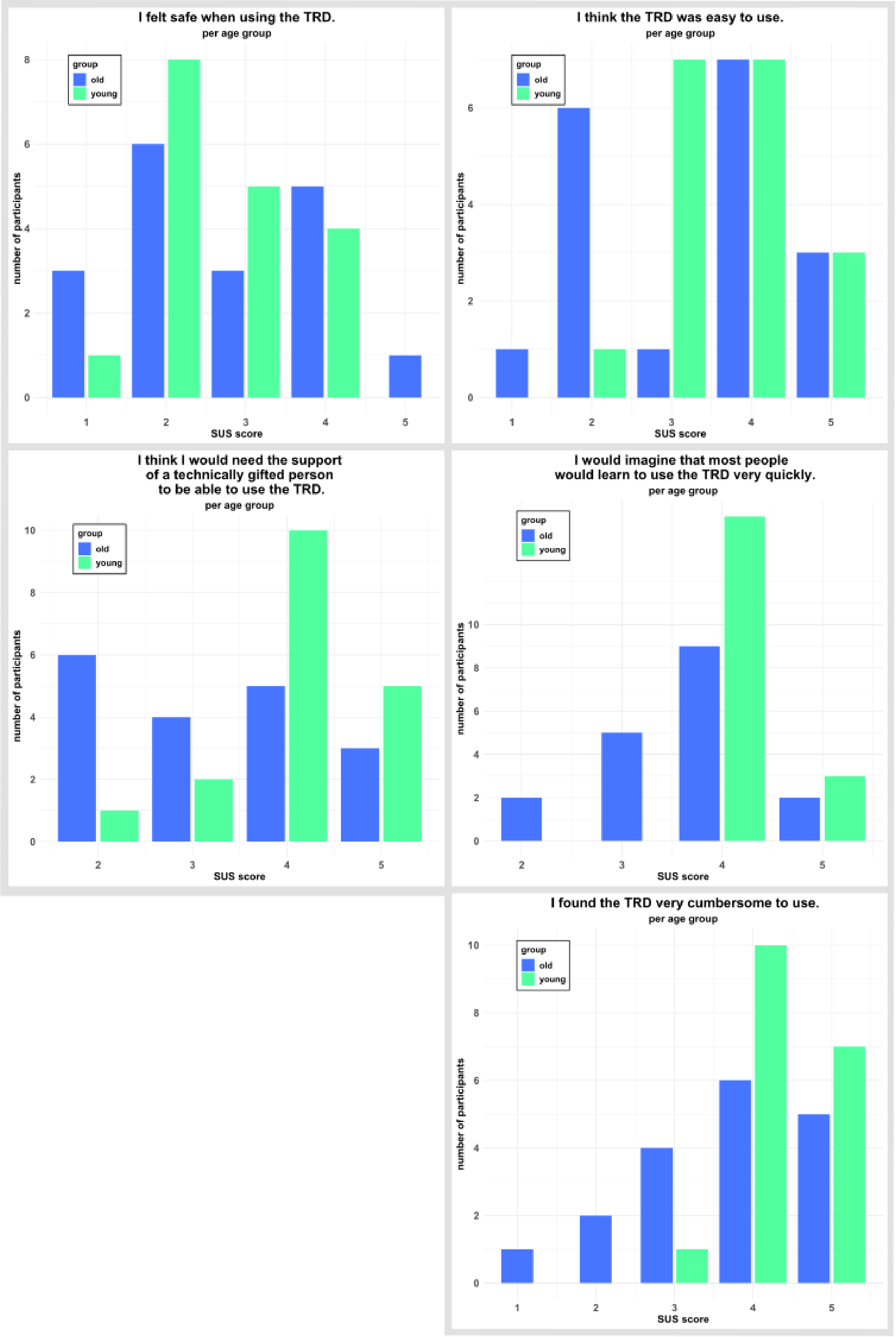
I felt safe when using the TRD.
per age group
group
old
young
number of participants
SUS score
I think the TRD was easy to use.
per age group
group
old
young
number of participants
SUS score
I think I would need the support of a technically gifted person to be able to use the TRD.
per age group
group
old
young
number of participants
SUS score
I would imagine that most people would learn to use the TRD very quickly.
per age group
group
old
young
number of participants
SUS score
I found the TRD very cumbersome to use.
per age group
group
old
young
number of participants
SUS score

**Figure S2. Individual item scores of the System Usability Scale (SUS).** The items were rated on a 5-point Likert scale (1 = “strongly disagree” to 5 = “strongly agree”). Bar plots represent the number of participants per possible item score, separated by age group.

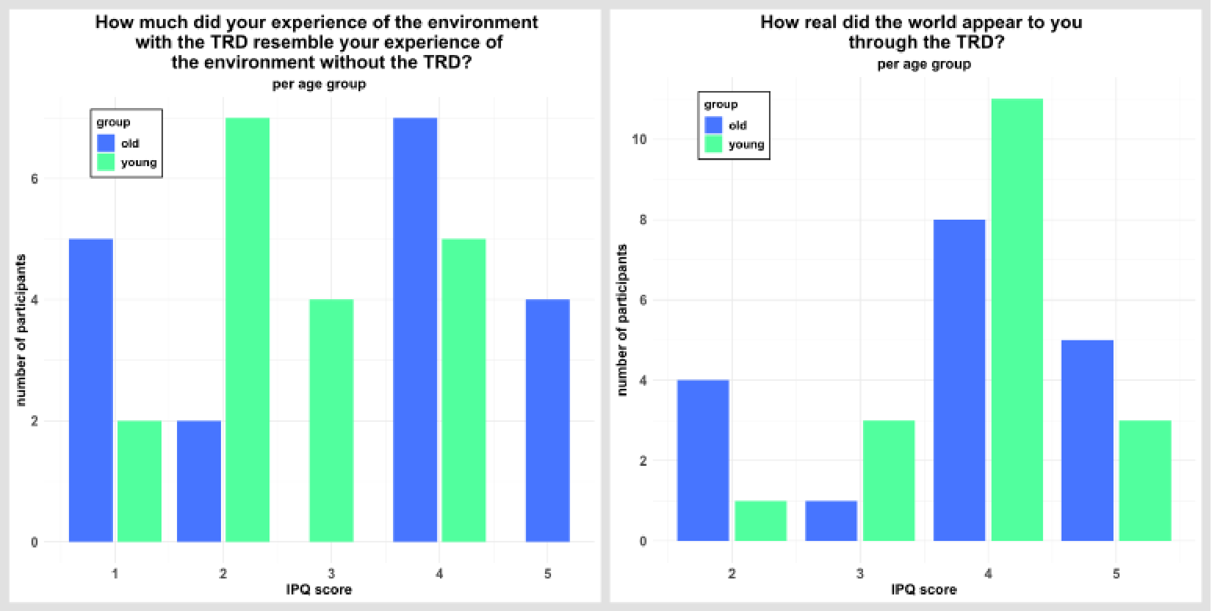


**Figure S3. Individual item scores of the iGroup Presence Questionnaire (IPQ).** The items were rated on a 5-point Likert scale (1 = “not real at all” to 5 = “perfectly real”). Bar plots represent the number of participants per possible item score, separated by age group.
